# Equitable Vaccine Access within an Age-Based Framework

**DOI:** 10.1101/2021.03.18.21253915

**Authors:** Alan B Cobo-Lewis

**Author notes:** **Corresponding Author:** Alan B. Cobo-Lewis, MS, PhD, 5717 Corbett Hall, Orono, ME 04469-5717, USA.

## Abstract

**Objectives:** Several states have adopted age-based prioritization for COVID-19 vaccine eligibility (also prioritizing teachers and child care workers) because it is simple (especially when age is quantized by decade) and age is strongly associated with COVID-19 mortality. But this approach raises equity concerns based in law and ethics. This study proposes data-driven solutions for equitable policy that retains the advantages of an age-based approach.

**Methods:** Using data from CDC and U.S. Census Bureau, I analyzed 507,227 COVID-19 deaths in the U.S. by age and race-ethnicity and compared the risk ratios to published data on risk ratios for other conditions.

**Results:** COVID-19 mortality in the U.S. rose 2.59-fold per decade of life [95% confidence interval (2.52, 2.66)]. Down syndrome, organ transplantation, and intellectual/developmental disability have risk ratios for COVID-19 mortality that exceed that.

**Conclusions:** People with specific conditions (including certain disabilities and certain medical conditions) associated with a risk ratio of 2.59 or 6.71 should be granted vaccine eligibility along with people 10 or 20 years older, respectively. Along with additional recommendations on data collection and reporting, this could help address equity within an age-based framework.

**3-Question Summary Box:**

- Some states have adopted age-only prioritization for COVID-19 vaccine access, quantized by decade of life, because it is simple, and age has a strong association with COVID-19 mortality. But this raises concerns based in law and ethics.
- This study quantifies the age effect and documents that COVID-19 mortality increases 2.59-fold per decade of life—a smaller increase than some conditions.
- People with conditions associated with at least 2.59-fold COVID-19 mortality should be granted vaccine eligibility along with people at least 10 years older.

---

Several U.S. states have moved to age-based prioritization for COVID-19 vaccine eligibility (modified to also prioritize teachers and child care workers, who President Biden has identified as a federal priority based on their role as essential workers for reopening schools rather than on increased COVID-19 risk *per se*). There is substantial variability in how states consider disability,^1^ but as of March 8, adults with high risk conditions are not prioritized in 12 states.^2^ Age-based prioritization is simple, especially when states quantize age by decade, opening vaccines to people at least in their 70s, then adding people in their 60s, etc. States have typically justified age-based prioritization by older people’s large risks of serious COVID-19 outcomes, including death, and by the need for simple, fast, and transparent systems^3,4^ But strictly age-based prioritization has come under assault as being unethical,^5^ and the U.S Department of Health and Human Services Office for Civil Rights has advised that, under the Affordable Care Act’s nondiscrimination provisions, a state or other entity is “only permitted to consider age as one factor as part of its overall decision-making.”^6^ Consequently, there are currently two complaints pending with the Office for Civil Rights challenging Connecticut’s age-based prioritization,^7,8^ and even as states do not all prioritize based on high-risk conditions, the federal government has very recently invited community health centers participating in the federal Health Center COVID-19 Vaccine Program to expand eligibility to persons with high-risk medical conditions, thus expanding access to the subset of COVID-19 vaccinations administered through this program.^9^

How to reconcile the large age-associated effects with ethical and legal demands for equity in jurisdictions that are prioritizing based solely on age? The answer is found in a proper quantification of the age-associated effects and a commitment to better data collection and reporting.

## Methods

To quantify the association of age (as well as race and ethnicity) with COVID-19 mortality, I downloaded publicly available data from the U.S. Centers for Disease Control and Prevention (CDC) and U.S. Census Bureau. Because the data are aggregated and de-identified, no IRB review was required.

Census data was from Vintage 2019 population estimates (the most recent available at time of analysis in March 2021). To analyze the overall increase of COVID-19 mortality by age, I downloaded COVID-19 data from https://data.cdc.gov/resource/9bhg-hcku.json. To analyze associations involving race or ethnicity, I downloaded additional COVID-19 data from https://data.cdc.gov/resource/ks3g-spdg.json. I constructed race and ethnicity groups to match those used in a previous study of racial and ethnic disparities:^10^ Hispanic, Non-Hispanic American Indian and Alaska Native, Non-Hispanic Asian or Pacific Islander (including Native Hawaiian), and Non-Hispanic Black. The three racial categories were for people reporting those races alone (but results were robust to including people of mixed race in those racial categories).

I conducted all analysis in conducted in R 4.0.4.^11^ COVID-19 mortality was calculated for 9 age categories that spanned a decade each (except for the youngest [0-14 years] and oldest [85+ years]). Using quasipoisson regression, I regressed COVID-19 deaths on age as well as jointly on age and racial-ethnic category, in both cases with an offset term for the logarithm of population. (This is equivalent to treating the expected value of the logarithm of COVID-19 mortality rate as a linear function of age or of age and racial-ethnic category.) In the regressions, I treated age as a linear predictor from 1 for the lowest group to 9 for the oldest group. In the regressions involving race and ethnicity, I analyzed racial-ethnic group with dummy coding, with overall COVID-19 mortality as the reference category.

I report all results with 95% confidence intervals.

To evaluate robustness to other methods of analysis, I also conducted logistic regressions of the COVID-19 mortality rates and linear regressions of the log-transformed COVID-19 mortality rates.

The full contents of the R code are included in supplementary material (in an R file). In order for the code to run for the first time, a census API must be obtained as described at https://api.census.gov/data/key_signup.html and installed using the census_api_key() function in the tidycensus R package.^12^ For a low-bandwidth analysis such as this, no API key is required from the CDC.

## Results

### Age

CDC data included data on 507,227 deaths from COVID-19 deaths in the U.S. Figure 1 plots COVID-19 mortality rate versus age, pooling across the entire population. Overall COVID-19 mortality in the U.S. increased by a factor of 2.59 (2.52, 2.66) per decade of life.

**Figure 1.**
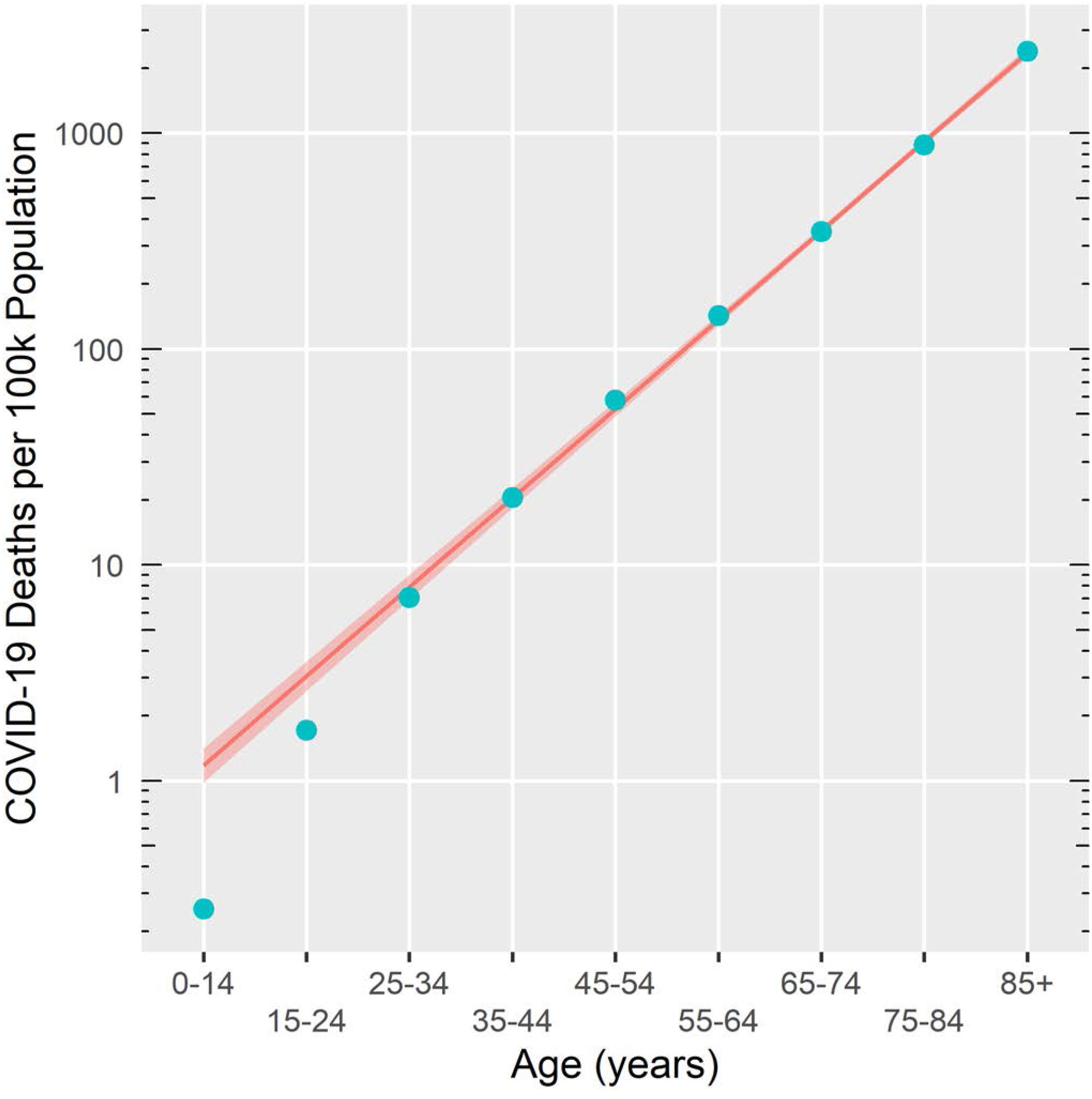
US Covid-19 mortality vs age for US population through March, 2021. Numerators for mortality rates are from CDC, and denominators are from U.S. Census Bureau. Line and 95% confidence interval reflect quasipoisson regression relating COVID-19 mortality to age and indicates that mortality increases by a factor of 2.59 per decade of life [95% confidence interval (2.52, 2.66)].

### Race and Ethnicity

Figure 2 presents the data for each racial-ethnic category in a separate panel, while also repeating the data and fit for the overall population in every panel. For each racial-ethnic group, the null hypothesis was that, adjusted for age, there was no difference between that group’s COVID-19 mortality and COVID-19 mortality in the overall population. The difference in intercept between racial-ethnic category and reference was largest for Non-Hispanic American Indian and Alaska Native (coefficient = 0.7534 [equivalent to risk ratio of exp(0.7534) = 2.12], *t* = 3.457, df = 39, *P* = 0.0013), then Hispanic (coefficient = 0.6622, risk ratio = 1.94, *t* = 11.169, df = 39, *P* < .0001), then Non-Hispanic Black (coefficient = 0.4456, risk ratio = 1.56, *t* = 6.806, df = 39, *P* < 0.0001). The difference in intercept was not significant between Non-Hispanic Asian or Pacific Islander and overall population (coefficient = - 0.1877, risk ratio = 0.83, t = -1.583, *P* = 0.12).

**Figure 2.**
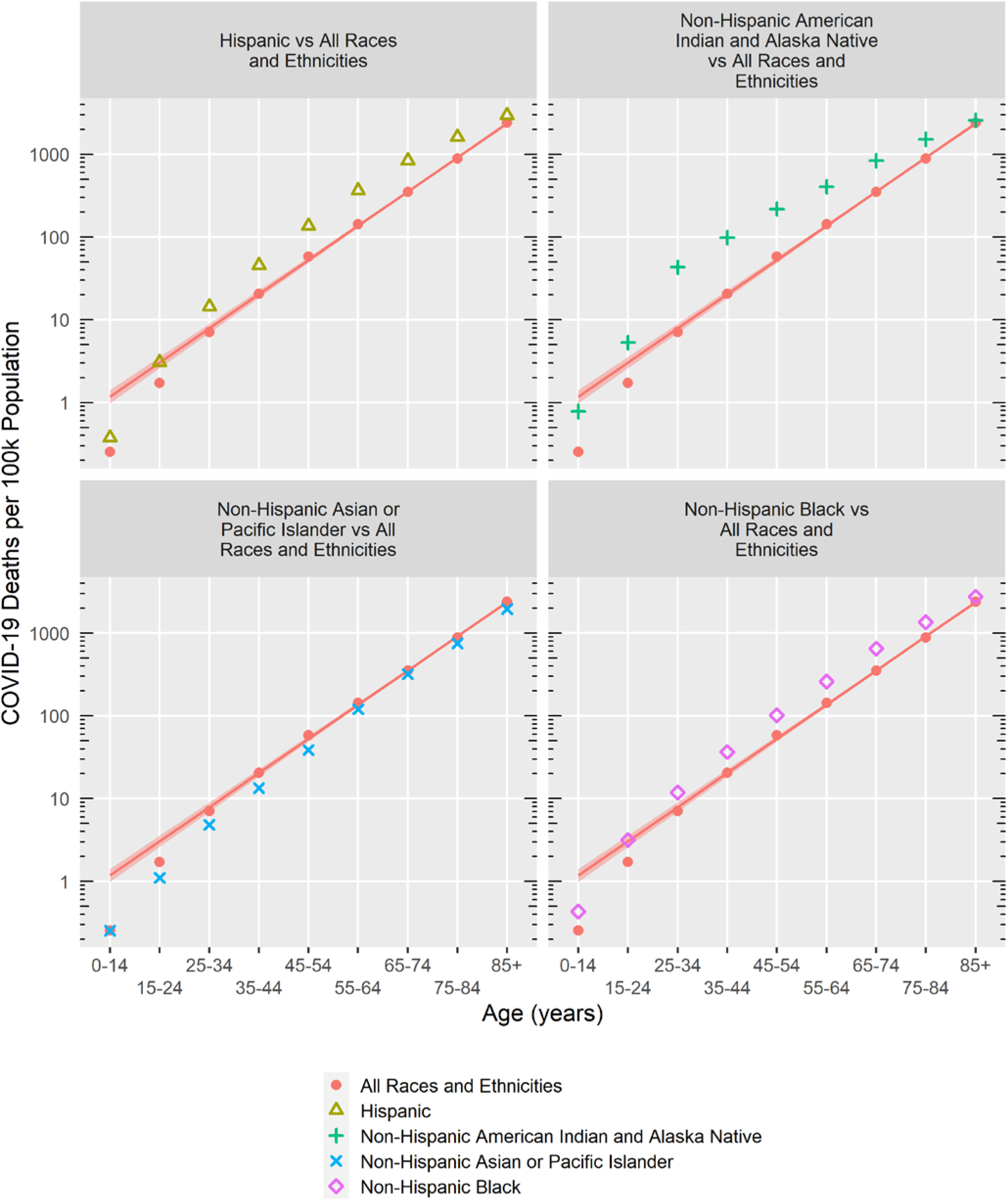
US Covid-19 mortality vs age, through March, 2021, for the entire US population and for four racial-ethnic groups. Numerators for mortality rates are from CDC, and denominators are from U.S. Census Bureau. Data and quasipoisson fit (including 95% confidence interval) for entire population are repeated in each panel, showing risk ratio for mortality increasing 2.59 per decade of life, [95% confidence interval (2.52, 2.66)].

COVID-19 mortality among racial and ethnic minorities also appeared especially elevated from levels in overall population in early adulthood through midlife, especially for Non-Hispanic American Indian and Alaska Native (note how the data in Figure 2 appear to follow a stronger quadratic trend in three of the four racial-ethnic groups than they do in the general population). Rather than pursuing curvilinear fits over a small number of age categories, for each racial-ethnic category, I instead compared the raw COVID-19 mortalities in age group directly to the corresponding values in the overall population (a model-free approach). The maximum risk ratio by age was similar to the regression-derived risk ratios for three racial-ethnic categories, but for the Non-Hispanic American Indian and Alaska Native, the risk ratio was as large as 6.08 (5.19, 7.12) for age 25-34, compared to a risk ratio of 2.12 (1.39, 3.26) for that racial-ethnic group’s regression-derived value.

### Sensitivity Analysis

When analyzed via quasilogistic regression instead of quasipoisson regression, the results are essentially the same (odds ratio of 2.60 per decade of life in quasilogistic regression vs risk ratio of 2.59 per decade of life in quasipoisson regression, for example). When log mortality rate is analyzed via linear regression, the age effect increases somewhat, to a risk ratio of 2.96 per decade of life, because linear regression increases the slope of the best-fitting line in Figures 1 and 2 to better fit the COVID-19 mortality in the two youngest age groups, whereas quasipoisson and quasilinear regression recognize the lower statistical reliability of those two data points.

## Discussion

Many potential risk factors elevate COVID-19 mortality by a factor less than the 2.59 risk ratio that people experience from a decade of life and might be excluded from prioritization with comparatively small effects on mortality equity (though a full consideration of equity would also consider effects beyond mortality). But a handful of conditions and demographic characteristics are associated with elevation in COVID-19 mortality that approaches or even exceeds 3-fold.

Figure 3 displays risk ratios deriving from the present study’s regressions, along with risk ratios from other studies for Down syndrome, intellectual or developmental disability, and organ transplant (except that, in one case,^13^ an odds ratio is displayed, but COVID-19 mortality is low enough that odds ratios and risk ratios can be treated similarly for those data).

**Figure 3.**
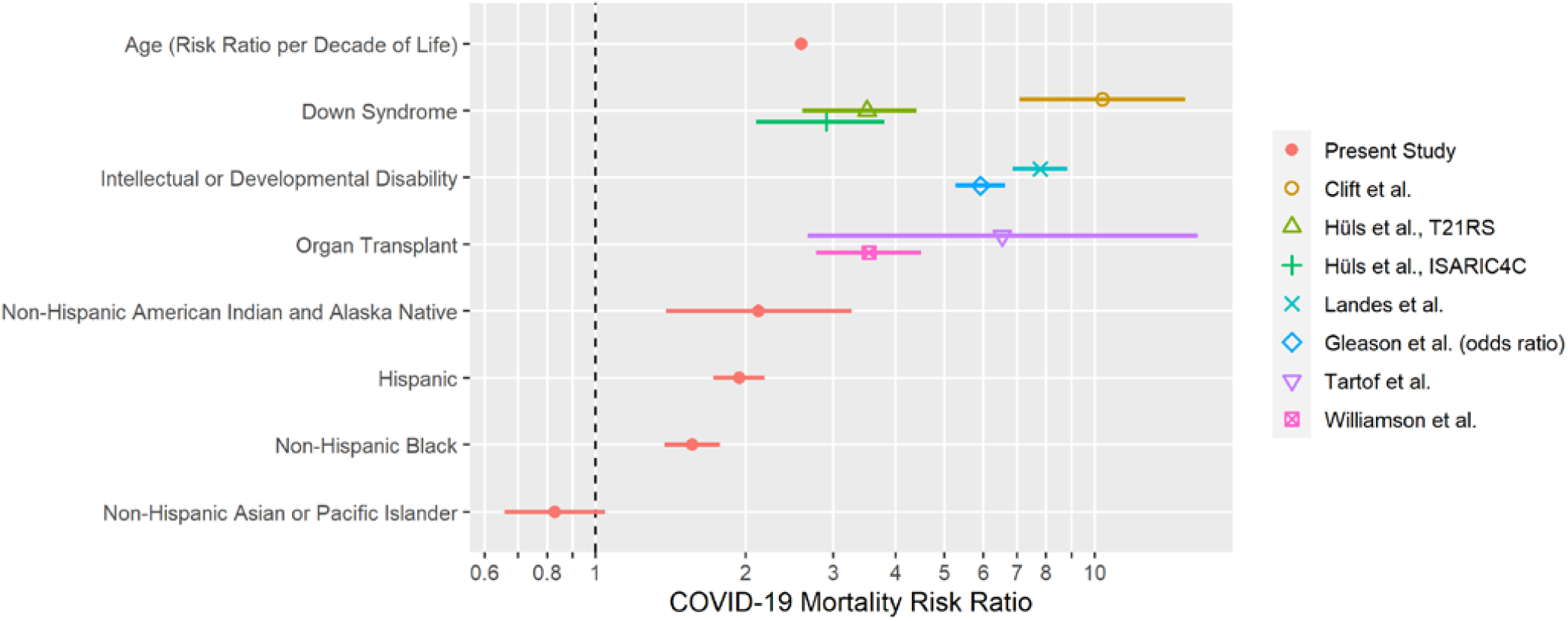
Covid-19 mortality risk ratios from present study for age and racial-ethnic category (US data through March, 2021) and from published studies for Down syndrome, intellectual or developmental disability, and organ transplant. Error bars indicate 95% confidence intervals.

Early in the pandemic, Down syndrome was reported to be associated with a 10-fold elevation in COVID-19 mortality,^14^ though an analysis of two more recent datasets has found elevation of 2.9- and 3.5-fold, respectively.^15^ Intellectual or developmental disability (a broader characteristic than Down syndrome) has been reported to be associated with an elevation in COVID-19 mortality of 5.9-fold^13^ to 7.8-fold.^16^ Organ transplantation has been reported to be associated with an elevation in COVID-19 mortality of 3.5-fold^17^ to 6.5-fold.^18^

Elevations in COVID-19 mortality by race and ethnicity are not as large in the present analysis as were found earlier in the pandemic,^10^ but they are still present. In particular, the point estimate is 2.9-fold for Non-Hispanic Native American Indian and Alaska Natives, though the risk ratio rose as high as 6.1-fold for 25-34-year-old Non-Hispanic Native American Indian and Alaska Natives.

## Conclusion

States that use a strictly age-based framework for prioritizing eligibility for vaccines typically justify that approach by noting the enormous risk ratio for COVID-19 mortality of the oldest groups versus young adults. For example, people at least 85 years old had a COVID-19 mortality of 2396.68 per 100k population, versus 7.08 per 100k among 25-34-year-olds—a risk ratio of 338.26. But this amounts to a risk ratio *per decade* of 338.26^1/6^ = 2.64, and states typically quantize age by decade—for example, opening up vaccines to 70-year-olds, then 60-years, etc. By calibrating potential risk factors for COVID-19 mortality against the risk ratio per decade (2.59 per decade when fitting all the data via quasipoisson regression), we can identify characteristics that should be considered for vaccine access even within an age-based framework. This analysis—and the data that informs such analyses—lead to specific policy prescriptions:

### Policy Prescription 1: Access to Vaccines

A 2.59-fold elevation in COVID-19 mortality is the same risk conveyed by being 10 years older, and a 6.70-fold elevation in COVID-19 mortality is the same risk conveyed by being 20 years older (because 2.59^2^ = 6.70). In order to begin to address equity, in any jurisdiction with an age-based framework for COVID-19 vaccine prioritization, people with conditions associated with a 2.59-fold elevation in COVID-19 mortality should become eligible for vaccines along with people 10 years older, and people with conditions associated with a 6.70-fold elevation in COVID-19 mortality should become eligible for vaccines along with people 20 year older. (If the 3-fold-per decade elevation from the linear regression were adopted then the policy prescription would be modified only slightly, so that people with conditions associated with a 3-fold increase in COVID-19 mortality should gain vaccine eligibility with people 10 years older and people with conditions associated with a 9-fold increase in COVID-19 mortality should gain vaccine eligibility.) For example, a 40-year-old with Down syndrome or organ transplantation should become eligible for COVID-19 vaccines at the same time as 50- or 60-year-olds from the general population. Important for speeding vaccine delivery, Down syndrome is easy to identify. This recommendation continues to be relevant in the U.S. until all adults become vaccine-eligible on May 1. It will also remain relevant several months or more into the future for countries that continue to experience vaccine shortage and that are considering an age-based eligibility framework.

### Policy Prescription 2: Access to Data

Down syndrome and organ transplantation are both found on the CDC’s list of medical conditions with sufficient evidence to conclude they put people at increased risk of severe illness from COVID-19.^19^ Molecular and genetic investigation has begun to suggest mechanisms that contribute to elevated COVID-19 risk in Down syndrome,^20^ and the immune suppressants associated with organ transplantation make the risk obvious. On the other hand, the strong association of intellectual or developmental disabilities overall with COVID-19 mortality may be at least partially explained by congregate living—90% of Americans with intellectual or developmental disabilities live alone, with a roommate, or with family caregivers,^21^ but people living in congregate settings are overrepresented in the published analyses about COVID-19 in people with intellectual or developmental disabilities, and some states that do not prioritize people with intellectual or developmental disabilities *per se* may prioritize a sub-population indirectly by prioritizing people living in congregate settings. It is therefore important to obtain unbiased estimates of the COVID-19 mortality risk among people with intellectual or developmental disabilities who live in less segregated and less congregate settings and to quantify the additional COVID-19 mortality risk that segregated and congregate settings may pose for this population. In addition, outside the U.S., data should be brought to bear to potentially modify the age-related 2.59-fold threshold according to local conditions of the pandemic.

In the U.S., state developmental disabilities agencies, for the most part, have COVID-19 data about people with disabilities—at least for people receiving services from state institutions or home- and community-based services funded through Medicaid—because COVID-19 cases and deaths constitute “critical incidents” that must be reported to the agency. But the data collection systems for people with intellectual developmental disabilities are largely segregated from the general public health data systems. Consequently, while COVID-19 dashboards, which are common throughout the U.S., track and report data by several risk factors (age and race among them), they rarely if ever report data on disability, even though states could link their public health and vaccine databases with databases supporting their developmental disabilities agencies or Medicaid agencies. This makes it impossible to assess the COVID-19 mortality associated with intellectual or developmental disability *per se*, and it also makes it impossible for the public to track vaccine progress in this marginalized population— important considerations even after vaccine eligibility is broadened to the general adult population. This must change.

## Data Availability

All data were downloaded from the CDC and the Census Bureau, where they are publicly available.

https://data.cdc.gov/resource/9bhg-hcku.json

https://data.cdc.gov/resource/ks3g-spdg.json

## Acknowledgments

Thanks to Angie Claussen for comments on a previous draft.

## Data and materials availability

All code is available in Supplementary Materials. All data are either included in the code or are publicly available from CDC and are accessed via the code.

## Supplementary Materials

Code

