## Supplementary material for "Equitable Vaccine Access within an Age-Based Framework": R code

# This code ran on 64-bit R 4.0.4 under Windows 10
library(data.table)
library(ggplot2)
library(tidycensus)
library(RSocrata)
library(ggeffects)
library(scales)

census_api_key(Sys.getenv('CENSUS_KEY')) # see http://api.census.gov/data/key_signup.html

# naming conventions:
#   data.table objects end with .dt
#   melted data.table objects end with .m.dt

# Analysis of Just Age and plotting Fig 1
# see CDC documentation at https://dev.socrata.com/foundry/data.cdc.gov/9bhg-hcku

# Do four reads, two from CDC and two from Census Bureau:
#
# Code run 2021mar05, at which time CDC data was updated 2021mar10 and had data through 2021mar06:
covid.by.age.dt <- data.table(read.socrata('https://data.cdc.gov/resource/9bhg-hcku.json'))
# Get vintage 2019 census data, because that's the latest available when code run:
pop.by.agegroup.us.dt <- data.table(get_estimates(geography='us', product='characteristics', breakdown='AGEGROUP', breakdown_labels=TRUE, year=2019))
# Code run 2021mar05, at which time CDC data was updated 2021mar10 and had data through 2021mar06
covid.by.age.race.eth.dt <- data.table(read.socrata('https://data.cdc.gov/resource/ks3g-spdg.json'))
# Get vintage 2019 census data, because that's the latest available when code run
pop.by.agegroup.race.eth.us.dt <- data.table(get_estimates(geography='us', product='characteristics', breakdown='AGEGROUP,RACE,HISP', breakdown_labels=TRUE, year=2019))


# Process the US Census data and bin age in a way to match up with CDC binning of age
pop.by.agegroup.us.cast.dt <- dcast(
  pop.by.agegroup.us.dt[grep('^Age', AGEGROUP)][
    ,.(NAME, pop=value,
       agegroup=AGEGROUP %>% gsub('Age ', 'age.', .) %>% gsub(' to ', '.', .) %>% gsub(' years', '', .) %>% gsub(' and older', '.and.older', .)
    )
  ],
  .~agegroup,
  value.var='pop'
)[ , .(
  age.0.14 = age.0.4 + age.5.9 + age.10.14,
  age.15.24 = age.15.19 + age.20.24,
  age.25.34 = age.25.29 + age.30.34,
  age.35.44 = age.35.39 + age.40.44,
  age.45.54 = age.45.49 + age.50.54,
  age.55.64 = age.55.59 + age.60.64,
  age.65.74 = age.65.69 + age.70.74,
  age.75.84 = age.75.79 + age.80.84,
  age.85.and.older = age.85.and.older
  )]
sum(pop.by.agegroup.us.cast.dt) # sums to 328M and change (2019 US population)
pop.by.agegroup.us.m.dt <- melt(pop.by.agegroup.us.cast.dt, value.name='pop', variable.name = 'age.group') # generates harmless warning about id.vars and measure.vars both being NULL

# Process the CDC covid data according to the same age bins as we used to process the census data
covid.by.age.us.dt <- covid.by.age.dt[
  data_as_of==max(data_as_of) & start_date==min(start_date) & end_date==max(end_date) & state=='United States' & sex=='All Sexes',
  .(covid_19_deaths),
  by=age_group
]
covid.by.age.us.dt[age_group %in% c('Under 1 year', '1-4 years', '5-14 years'), age.group := 'age.0.14'] # age_group was in CDC dataset but we'll call our new variable age.group
covid.by.age.us.dt[age_group != '18-29 years' & age_group != '30-39 years' & age_group != '40-49 years' & age_group != '50-64 years' & grepl('^[0-9][0-9]', age_group), age.group := age_group %>% gsub('^', 'age.', .) %>% gsub('-', '.', .) %>% gsub(' years and over', '.and.older', .) %>% gsub(' years', '', .)]
covid.by.age.group.us.dt <- covid.by.age.us.dt[!is.na(age.group), .(covid_19_deaths = sum(as.numeric(covid_19_deaths))), by=age.group] # need as.numeric() because covid_19_deaths variable was a string


# Merge covid data with census data
covid.and.pop.by.age.group.us.dt <- covid.by.age.group.us.dt[pop.by.agegroup.us.m.dt, on='age.group']
covid.and.pop.by.age.group.us.per.100k.dt <- covid.and.pop.by.age.group.us.dt[ , .(covid_19_deaths_per_100k=1e5*covid_19_deaths/pop, number.of.covid.19.deaths=as.integer(covid_19_deaths), pop=as.integer(pop)), by='age.group']

# Code run 2021mar05, at which time CDC data was updated 2021mar10 and had data through 2021mar06, with 507,227 covid-19 deaths:
covid.and.pop.by.age.group.us.per.100k.dt[,sum(number.of.covid.19.deaths)]

# The fit that is presented in the paper is quasipoisson, but code is also presented for sensitivity checks (quasilogit and linear fits):
#   (All three of these models have linear trend through age.group, treating youngest and oldest groups like the others)
fit.quasipoisson <- glm(
  number.of.covid.19.deaths ~ offset(log(pop)) + as.numeric(factor(age.group)),
  family=quasipoisson,
  data=covid.and.pop.by.age.group.us.per.100k.dt
)
fit.quasilogit <- glm(
  cbind(number.of.covid.19.deaths, pop-number.of.covid.19.deaths) ~ as.numeric(factor(age.group)),
  family=quasibinomial,
  data=covid.and.pop.by.age.group.us.per.100k.dt
)
fit.linear <- lm(log10(covid_19_deaths_per_100k) ~ as.numeric(factor(age.group)), data=covid.and.pop.by.age.group.us.per.100k.dt)
summary(fit.quasipoisson)
exp(coef(fit.quasipoisson)) # risk ratio = 2.59-fold per decade
exp(coef(fit.quasilogit)) # odds ratio = 2.60-fold per decade
10^coef(fit.linear) # risk ratio = 2.96-fold per decade
# confidence intervals for the RRs a
exp(confint(fit.quasipoisson))
exp(confint(fit.quasilogit))
10^(confint(fit.linear))


fit.us.preds.quasipoisson <- as.data.table(ggemmeans(fit.quasipoisson, terms='age.group', condition=c(pop=1e5)))

windows()
ggplot(
  covid.and.pop.by.age.group.us.per.100k.dt,
  aes(
    x=age.group,
    y=covid_19_deaths_per_100k)
  ) +
  xlab('Age (years)') +
  geom_ribbon(group=1, data=fit.us.preds.quasipoisson, aes(fill='fit', x=x, y=NULL, ymin=conf.low, ymax=conf.high), alpha=.4) +
  geom_line(group=1, size=1, data=fit.us.preds.quasipoisson, aes(color='fit', x=x, y=predicted)) +
  geom_point(size=4, aes(color='US CDC Data')) +
  scale_y_log10(minor_breaks=NULL) +
  annotation_logticks(sides='lr') +
  scale_fill_discrete(guide='none') +
  scale_x_discrete(
    labels=function(x) x %>% gsub('age[.]85[.]and[.]older', '85+', .) %>% gsub('age[.]', '', .) %>% gsub('[.]', '-', .),
    guide=guide_axis(n.dodge=2)
  ) + 
  ylab('COVID-19 Deaths per 100k Population') +
  theme_gray(base_size=18) + theme(legend.position='none', legend.direction='vertical', legend.title=element_blank(), axis.ticks.y=element_blank())
ggsave('PHR-Fig-1.png')
ggsave('PHR-Fig-1.eps')

###################

# Analysis of Age and Race-Ethnicity, plotting Figure 2, plotting Figure 2

# use the four marginalized racial-ethnic groups of Bassett et al. (2020) in population calculations
pop.by.agegroup.raceeth4.us.dt <- copy(pop.by.agegroup.race.eth.us.dt)
pop.by.agegroup.raceeth4.us.dt[RACE=='Black alone' & HISP=='Non-Hispanic' , raceeth4 := 'Non-Hispanic Black']
pop.by.agegroup.raceeth4.us.dt[RACE=='All races' & HISP=='Hispanic', raceeth4 := 'Hispanic']
pop.by.agegroup.raceeth4.us.dt[RACE=='American Indian and Alaska Native alone' & HISP=='Non-Hispanic', raceeth4 := 'Non-Hispanic American Indian and Alaska Native']
pop.by.agegroup.raceeth4.us.dt[RACE=='Asian alone' & HISP=='Non-Hispanic', raceeth4 := 'Non-Hispanic Asian or Pacific Islander']
pop.by.agegroup.raceeth4.us.dt[RACE=='Native Hawaiian and Other Pacific Islander alone' & HISP=='Non-Hispanic', raceeth4 := 'Non-Hispanic Asian or Pacific Islander']
pop.by.agegroup.raceeth4.us.dt <- pop.by.agegroup.raceeth4.us.dt[!is.na(raceeth4) & AGEGROUP != 'Median age']
pop.by.agegroup.raceeth4.us.dt <- pop.by.agegroup.raceeth4.us.dt[ , .(value=sum(value)), by=.(AGEGROUP, raceeth4)] # Following Bassett et al., combine NH Asian with NH Pacific Islander

# use the same age groups in population table as are used in the CDC's covid dataset
pop.by.agegroup.raceeth4.us.cast.dt <- dcast(
  pop.by.agegroup.raceeth4.us.dt[grep('^Age', AGEGROUP)][
    ,.(raceeth4, pop=value,
       agegroup=AGEGROUP %>% gsub('Age ', 'age.', .) %>% gsub(' to ', '.', .) %>% gsub(' years', '', .) %>% gsub(' and older', '.and.older', .)
    )
    ],
  raceeth4~agegroup,
  value.var='pop'
)[ , .(raceeth4,
  age.0.14 = age.0.4 + age.5.9 + age.10.14,
  age.15.24 = age.15.19 + age.20.24,
  age.25.34 = age.25.29 + age.30.34,
  age.35.44 = age.35.39 + age.40.44,
  age.45.54 = age.45.49 + age.50.54,
  age.55.64 = age.55.59 + age.60.64,
  age.65.74 = age.65.69 + age.70.74,
  age.75.84 = age.75.79 + age.80.84,
  age.85.and.older = age.85.and.older
)]
pop.by.agegroup.raceeth4.us.m.dt <- melt(pop.by.agegroup.raceeth4.us.cast.dt, value.name='pop', variable.name='age.group') # generates harmless warning about id.vars and measure.vars both being NULL

# now use the same four margialized racial-ethnic groups of Bassett et al (2020) in processing the CDC data
covid.by.age.raceeth4.us.dt <- covid.by.age.race.eth.dt[
  data_as_of==max(data_as_of) & end_week==max(end_week) & state=='United States' & race_and_hispanic_origin %in% c('Non-Hispanic Black', 'Hispanic', 'Non-Hispanic American Indian or Alaska Native', 'Non-Hispanic Asian', 'Non-Hispanic Native Hawaiian or Other Pacific Islander'),
  .(race_and_hispanic_origin, covid_19_deaths, total_deaths, pneumonia_deaths, pneumonia_and_covid_19_deaths),
  by=age_group_new
  ]
covid.by.age.raceeth4.us.dt[race_and_hispanic_origin=='Non-Hispanic Black', raceeth4 := 'Non-Hispanic Black']
covid.by.age.raceeth4.us.dt[race_and_hispanic_origin=='Hispanic', raceeth4 := 'Hispanic']
covid.by.age.raceeth4.us.dt[race_and_hispanic_origin=='Non-Hispanic American Indian or Alaska Native', raceeth4 := 'Non-Hispanic American Indian and Alaska Native']
covid.by.age.raceeth4.us.dt[race_and_hispanic_origin=='Non-Hispanic Asian', raceeth4 := 'Non-Hispanic Asian or Pacific Islander']
covid.by.age.raceeth4.us.dt[race_and_hispanic_origin=='Non-Hispanic Native Hawaiian or Other Pacific Islander', raceeth4 := 'Non-Hispanic Asian or Pacific Islander']
covid.by.age.raceeth4.us.dt <- covid.by.age.raceeth4.us.dt[ , .(covid_19_deaths=sum(as.numeric(covid_19_deaths))), by=.(age_group_new, raceeth4)]

covid.by.age.raceeth4.us.dt[age_group_new %in% c('Under 1 year', '1-4 years', '5-14 years'), age.group := 'age.0.14']
covid.by.age.raceeth4.us.dt[age_group_new != '18-29 years' & age_group_new != '30-49 years' & age_group_new != '30-39 years' & age_group_new != '40-49 years' & age_group_new != '50-64 years' & grepl('^[0-9][0-9]', age_group_new), age.group := age_group_new %>% gsub('^', 'age.', .) %>% gsub('-', '.', .) %>% gsub(' years and over', '.and.older', .) %>% gsub(' years', '', .)]
covid.by.age.group.raceeth4.us.dt <- covid.by.age.raceeth4.us.dt[!is.na(age.group), .(covid_19_deaths=sum(as.numeric(covid_19_deaths))), by=.(raceeth4,age.group)]

# Merge covid data with census data
covid.and.pop.by.age.group.raceeth4.us.dt <- covid.by.age.group.raceeth4.us.dt[pop.by.agegroup.raceeth4.us.m.dt, on=c('raceeth4', 'age.group')]
covid.and.pop.by.age.group.raceeth4.us.per.100k.dt <- covid.and.pop.by.age.group.raceeth4.us.dt[ , .(covid_19_deaths_per_100k=1e5*covid_19_deaths/pop, number.of.covid.19.deaths=as.integer(covid_19_deaths), pop=as.integer(pop)), by=.(raceeth4, age.group)]

# combine covid-19 mortality by age group for the 4 marginalized racial-ethnic groups with covid-19 mortlity overall and calculate the ratio of covid-19 mortality rate for each combination of age group and racial-ethnic group
covid.and.pop.by.age.group.raceeth4.us.per.100k.vs.allraceeth.dt <- covid.and.pop.by.age.group.raceeth4.us.per.100k.dt[
  covid.and.pop.by.age.group.us.per.100k.dt[ , .(age.group, covid_19_deaths_per_100k.allracesandeth=covid_19_deaths_per_100k, number.of.covid.19.deaths.allracesandeth=number.of.covid.19.deaths, pop.allracesandeth=pop)],
  on='age.group'
][,.(raceeth4, age.group, covid_19_deaths_per_100k, covid_19_deaths_per_100k.allracesandeth, c19.death.elevation=covid_19_deaths_per_100k/covid_19_deaths_per_100k.allracesandeth, number.of.covid.19.deaths, number.of.covid.19.deaths.allracesandeth, pop, pop.allracesandeth)]

# model-free risk ratios and their CIs:
covid.and.pop.by.age.group.raceeth4.us.per.100k.vs.allraceeth.dt[ , `:=`(
  ln.rr.ci.half.width = # half-width of confidence interval for ln(risk ratio)
  qnorm(.975)*sqrt(
    ( (pop-number.of.covid.19.deaths)/number.of.covid.19.deaths / pop ) +
    ( (pop.allracesandeth-number.of.covid.19.deaths.allracesandeth)/number.of.covid.19.deaths.allracesandeth / pop.allracesandeth )
  )
)]
covid.and.pop.by.age.group.raceeth4.us.per.100k.vs.allraceeth.dt[ , `:=`(
  c19.death.elevation.lo=exp(log(c19.death.elevation)-ln.rr.ci.half.width), c19.death.elevation.hi=exp(log(c19.death.elevation)+ln.rr.ci.half.width)
)]
covid.and.pop.by.age.group.raceeth4.us.per.100k.vs.allraceeth.dt[c19.death.elevation==max(c19.death.elevation)] # pick out the joint combination of age.group and racial-ethnic group with the largest risk ratio relative to same age.group for overall population

raceeth5.levs <- c('All Races and Ethnicities', covid.and.pop.by.age.group.raceeth4.us.per.100k.vs.allraceeth.dt[,sort(unique(raceeth4))]) # raceeth5 includes "5th level" of overall (that actually includes more than just these four categories)
windows(10,12)
ggplot(covid.and.pop.by.age.group.raceeth4.us.per.100k.vs.allraceeth.dt, aes(age.group, covid_19_deaths_per_100k)) +
  facet_wrap(~paste(raceeth4, 'vs All Races and Ethnicities'), labeller=label_wrap_gen()) +
  geom_ribbon(group=1, data=fit.us.preds.quasipoisson[, cbind(.SD, raceeth5='All Races and Ethnicities')], aes(fill='fit', x=x, y=NULL, ymin=conf.low, ymax=conf.high), alpha=.4, show.legend=FALSE) +
  geom_line(group=1, size=1, data=fit.us.preds.quasipoisson[, cbind(.SD, raceeth5='All Races and Ethnicities')], aes(color='All Races and Ethnicities', shape='All Races and Ethnicities', x=x, y=predicted), show.legend=FALSE) +
  geom_point(size=3, aes(color='All Races and Ethnicities', shape='All Races and Ethnicities'), data=covid.and.pop.by.age.group.us.per.100k.dt) +
  geom_point(size=2, stroke=1.5, aes(color=raceeth4, shape=raceeth4)) +
  scale_color_discrete(name='group', labels=raceeth5.levs) +
  scale_shape_manual(name='group', labels=raceeth5.levs, values=c(16, 2:5)) +
  scale_y_log10(minor_breaks=NULL) +
  annotation_logticks(sides='lr') +
  scale_fill_discrete(guide='none') +
  scale_x_discrete(
    labels=function(x) x %>% gsub('age[.]85[.]and[.]older', '85+', .) %>% gsub('age[.]', '', .) %>% gsub('[.]', '-', .),
    guide=guide_axis(n.dodge=2)
  ) + 
  ylab('COVID-19 Deaths per 100k Population') +
  xlab('Age (years)') +
  theme_gray(base_size=16) + theme(legend.position='bottom', legend.direction='vertical', legend.title=element_blank(), axis.ticks.y=element_blank())
ggsave('PHR-Fig-2.png')
ggsave('PHR-Fig-2.eps')

# The fit that is presented in the paper is quasipoisson, but code is also presented for sensitivity checks (quasilogit and linear fits):
fit.raceeth5.quasipoisson <- glm(
  number.of.covid.19.deaths ~ offset(log(pop)) + as.numeric(factor(age.group)) + raceeth5,
  family=quasipoisson,
  data=rbindlist(list(
    covid.and.pop.by.age.group.us.per.100k.dt[, .(age.group, number.of.covid.19.deaths, pop, covid_19_deaths_per_100k, raceeth5='All Races and Ethnicities')],
    covid.and.pop.by.age.group.raceeth4.us.per.100k.dt[, .(age.group, number.of.covid.19.deaths, pop, covid_19_deaths_per_100k, raceeth5=raceeth4)]
  ))
)
fit.raceeth5.quasilogit <- glm(
  cbind(number.of.covid.19.deaths, pop-number.of.covid.19.deaths) ~ as.numeric(factor(age.group)) + raceeth5,
  family=quasibinomial,
  data=rbindlist(list(
    covid.and.pop.by.age.group.us.per.100k.dt[, .(age.group, number.of.covid.19.deaths, pop, covid_19_deaths_per_100k, raceeth5='All Races and Ethnicities')],
    covid.and.pop.by.age.group.raceeth4.us.per.100k.dt[, .(age.group, number.of.covid.19.deaths, pop, covid_19_deaths_per_100k, raceeth5=raceeth4)]
  ))
)
fit.raceeth5.linear <- lm(
  log10(number.of.covid.19.deaths) ~ as.numeric(factor(age.group)) + raceeth5,
  data=rbindlist(list(
    covid.and.pop.by.age.group.us.per.100k.dt[, .(age.group, number.of.covid.19.deaths, pop, covid_19_deaths_per_100k, raceeth5='All Races and Ethnicities')],
    covid.and.pop.by.age.group.raceeth4.us.per.100k.dt[, .(age.group, number.of.covid.19.deaths, pop, covid_19_deaths_per_100k, raceeth5=raceeth4)]
  ))
)
summary(fit.raceeth5.quasipoisson) # for t ratios etc. for Results section
exp(coef(fit.raceeth5.quasipoisson))
exp(confint(fit.raceeth5.quasipoisson))
exp(coef(fit.raceeth5.quasilogit))
exp(confint(fit.raceeth5.quasilogit))
10^coef(fit.raceeth5.linear)
10^confint(fit.raceeth5.linear)

################

summary.effects.quasipoisson.dt <- rbindlist(list(
  as.data.table(summary(fit.raceeth5.quasipoisson)$coefficients, keep.rownames = TRUE)[-(1:2) , .(group='Race & Ethnicity', label='Present Study', rn = rn %>% gsub('raceeth5', '', .), y=exp(Estimate), y.lo=exp(Estimate-qnorm(.975)*`Std. Error`), y.hi=exp(Estimate+qnorm(.975)*`Std. Error`))],
  data.table(
    group=c('Organ Transplant', 'Organ Transplant', 'Down Syndrome', 'Down Syndrome', 'Down Syndrome', 'Intellectual or Developmental Disability', 'Intellectual or Developmental Disability'),
    label=c('Williamson et al.', 'Tartof et al.', 'H�ls et al., T21RS', 'H�ls et al., ISARIC4C', 'Clift et al.', 'Landes et al.', 'Gleason et al. (odds ratio)'),
    rn=c('Organ Transplant', 'Organ Transplant', 'Down Syndrome', 'Down Syndrome', 'Down Syndrome', 'Intellectual or Developmental Disability', 'Intellectual or Developmental Disability'),
    y=c(3.53, 6.54, 3.5, 2.9, 10.39, 7.781, 5.909),
    y.lo=c(2.77, 2.66, 2.6, 2.1, 7.08, 6.856, 5.277),
    y.hi=c(4.49, 16.12, 4.4, 3.8, 15.23, 8.832, 6.617)
  )
), use.names=TRUE, fill=TRUE)
rn.levels <- c(summary.effects.quasipoisson.dt[group=='Race & Ethnicity', levels(reorder(rn, y))], 'Organ Transplant', 'Intellectual or Developmental Disability', 'Down Syndrome', 'Age (Risk Ratio per Decade of Life)')
label.levels <- c('Present Study', 'Clift et al.', 'H�ls et al., T21RS', 'H�ls et al., ISARIC4C', 'Landes et al.', 'Gleason et al. (odds ratio)', 'Tartof et al.', 'Williamson et al.')
decade.effect.quasipoisson.dt <- as.data.table(summary(fit.quasipoisson)$coefficients, keep.rownames = TRUE)[-1 , .(group=NULL, label='Present Study', rn ='Age (Risk Ratio per Decade of Life)', y=exp(Estimate), y.lo=exp(Estimate-qnorm(.975)*`Std. Error`), y.hi=exp(Estimate+qnorm(.975)*`Std. Error`))]
windows(10,4)
summary.and.decade.effects.quasipoisson.dt <- rbindlist(list(decade.effect.quasipoisson.dt, summary.effects.quasipoisson.dt), use.names=TRUE, fill=TRUE)
ggplot(summary.and.decade.effects.quasipoisson.dt, aes(y=y, ymin=y.lo, ymax=y.hi, x=rn, shape=factor(label, levels=label.levels), color=factor(label, levels=label.levels))) +
  geom_hline(yintercept=1, linetype='dashed') +
  geom_point(stroke=1, size=2, na.rm=TRUE, position=position_dodge(width=-.5)) +
  geom_linerange(size=1, show.legend=FALSE, na.rm=TRUE, position=position_dodge(width=-.5)) +
  scale_shape_manual(name='label', labels=label.levels, values=c(16, 1:9)) +
  scale_color_discrete(name='label', labels=label.levels) +
  coord_flip() +
  scale_x_discrete(limits=rn.levels) +
  scale_y_log10(minor_breaks=NULL, breaks=c(.1*(1:9), 1:9, 10*(1:9)), labels=c(.1, .2, .3, .4, '', .6, '', .8, '', 1:8, '', 10*(1:9))) +
  ylab('COVID-19 Mortality Risk Ratio') +
  theme_gray(base_size=12) + theme(axis.title.y=element_blank(), legend.title=element_blank())
ggsave('PHR-Fig-3.png')
ggsave('PHR-Fig-3.eps')
